# White Matter Abnormalities in Bipolar II and Unipolar Depression – Evidence from Fixel-Based Analysis

**DOI:** 10.64898/2026.01.22.26344600

**Authors:** Idy W. Y. Chou, Anna Manelis, Holly A. Swartz, Owen N. W. Leung, Mary L. Phillips, Suzanne H. W. So, Winnie C. W. Chu, Hanna Lu, Linda C. W. Lam, Arthur D. P. Mak

**Affiliations:** Department of Psychiatry, The Chinese University of Hong Kong; Department of Psychiatry, University of Pittsburgh; Department of Psychology, The Chinese University of Hong Kong; Department of Imaging & Interventional Radiology, The Chinese University of Hong Kong

**Author notes:** **Corresponding author:** Chou, Idy W. Y. The Chinese University of Hong Kong, G/F Multicentre, Tai Po Hospital, Tai Po, Hong Kong.

**Keywords:** Bipolar disorder, Depressive disorders, White matter abnormalities, Diffusion-weighted imaging, Fixel-based analysis, Fibre density, FDC

## Abstract

**Background:** Challenges in correctly identifying bipolar II disorder (BD-II) during depressive states have led to poor clinical outcomes. BD-II-specific imaging investigations are lacking. This study addresses current knowledge gaps by comparing white matter (WM) integrity in BD-II and unipolar depression (UD) using fixel-based analysis.

**Method:** Fibre density (FD), fibre cross-section (FC), and the combined measure (FDC) within 72 WM tracts were compared among 33 individuals with BD-II, 50 with UD, and 51 healthy controls (HC). The effects of illness characteristics on FBA correlates were also examined. Sensitivity analyses compared these measures among unmedicated participants to check whether medication status affects the results.

**Results:** Participants with BD-II and UD showed reduced FD in the left parieto-occipito-pontine (POPT) and striato-occipital (ST-OCC) tracts. Compared to UD, BD-II was associated with lower FD in the left arcuate fascicle (AF) and bilateral superior longitudinal fasciculi I and II (SLF-I and II). In BD-II, illness duration negatively correlated with FD in left AF, left POPT, and right ST-OCC, while the number of lifetime BD-II depressive episodes positively correlated with FDC in left SLF-I. Group differences were significant but less pronounced in unmedicated participants.

**Conclusions:** Our findings demonstrate shared and distinct WM abnormalities in tracts involved in visuomotor and executive processes in BD-II and UD, with BD-II exhibiting more extensive alterations. With BD-II, but not UD, longer illness duration was linked to lower FD, while depression recurrence was associated with higher FDC, suggesting potential degenerative and compensatory neurobiological mechanisms. Longitudinal studies should investigate the joint trajectories of symptomatology and WM alterations.

## 1. Introduction

Bipolar II disorder (BD-II) is often misdiagnosed as unipolar depression (UD) owing to the predominance of depressive over hypomanic episodes, shared diagnostic criteria for depressive episodes, and egosyntonic nature of hypomania (Forte et al., 2015; Judd et al., 2003; Swartz & Suppes, 2023), leading to delays in diagnosis and treatment (Keramatian et al., 2022). Individuals with BD-II experience disabling depression with higher recurrence risk compared to those with UD, and, unlike the latter, mood stabiliser but not antidepressant medications are the most appropriate treatment (Kennedy et al., 2016; Maina et al., 2007; Yatham et al., 2018). Investigating the neurobiological mechanisms of BD-II, particularly the differences from UD, may offer a promising avenue for improving diagnosis and developing targeted interventions.

White matter (WM) consists of myelinated axons ensuring communication across brain regions. Growing evidence indicates impaired energy metabolism in BD, whereby increased oxidative stress triggers neuroinflammation, demyelination, and axonal degeneration (Berk et al., 2011; Dean & Keshavan, 2017; Scaini et al., 2020). Investigation of the WM alterations in mood disorders can shed light into the underlying neurobiological mechanisms. Indeed, WM abnormalities are well-documented in mood disorders, with BD showing more extensive disruptions than UD (Bauer et al., 2016; Koshiyama et al., 2020). Diffusion tensor imaging (DTI) studies consistently report reduced fractional anisotropy (FA) in BD across major association tracts – the inferior fronto-occipital fasciculus (IFO), inferior and superior longitudinal fasciculi (ILF, SLF), uncinate fasciculus (UF), cingulum (CG), and corpus callosum (CC) – suggesting widespread microstructural compromise (Caseras et al., 2013; Foley et al., 2018; Ha et al., 2009; Yang et al., 2019). These tracts support emotion regulation, executive control, and interhemispheric communication (Conner et al., 2018; Herbet et al., 2018; Kamali et al., 2014; Martino et al., 2010; Philippi et al., 2009; Von Der Heide et al., 2013). Their disruption may underlie bipolar-specific affective, cognitive, and sensory dysfunctions (Elliott et al., 2011; Engel-Yeger et al., 2016; Masuda et al., 2020; Panchal et al., 2019; Shaffer Jr et al., 2018). A large-scale mega-analysis identified the CG and CC as showing the most pronounced FA reductions in BD (Favre et al., 2019).

Most WM studies on BD have focused on BD-I or BD-I/BD-II mixed subtypes. Only two studies directly compared WM integrity between BD-II and UD. Manelis et al. (2021) found reduced FA in BD-II relative to UD in the left arcuate fascicle (AF), whereas Mak et al. (2021) reported no significant FA differences between BD-II and UD in medication-naïve participants, thus raising a question of how exposure to psychotropic medications contributes to WM integrity alterations, with existing evidence pointing to both protective (Abramovic et al., 2018; Benedetti et al., 2011; Favre et al., 2019) and detrimental (Mamah et al., 2019) effects. Furthermore, differences in scanning parameters – Manelis et al. (2021) used diffusion gradients with b=2000s/mm^2^ along 150 directions, whereas Mak et al. (2021) employed b=1000s/mm^2^ along 32 directions – may have caused differing sensitivity to microstructural differences.

Another challenge in WM investigation arises from the technical limitation of DTI in resolving crossing fibres, causing inaccurate tract reconstruction (Mangin et al., 2013). Fixel-based analysis (FBA) is an advanced analysis technique which overcomes this limitation by estimating fibre orientation distributions (FODs) of multiple fibre populations per voxel, represented as “fixels” (Raffelt et al., 2017; Tournier et al., 2008). Several metrics can be derived, including fibre density (FD) that reflects intra-axonal volume, with decreases indicating axonal loss/degeneration; fibre cross-section (FC) that captures macroscopic WM morphology, where decreases reflect atrophy or demyelination; and the combined measure of FD and FC (FDC) which summarizes both measures and represents overall capacity of fibre bundles in transmitting information (Dhollander et al., 2021; Raffelt et al., 2012; Raffelt et al., 2017). Only two studies have used FBA to investigate WM abnormalities in mood disorders. Lyon et al. (2019) found in UD decreased FD in CC and decreased FC in the anterior limb of the internal capsule. Also, higher FC in a subpart of CC (tapetum) predicted depression remission. Manelis et al. (2025) reported reduced FD in both BD and UD in the splenium of CC, left optic radiation (OR), and left striato-occipital tract (ST-OCC). Relative to UD, BD had decreased FD in the right SLF, right UF, and left thalamo-occipital tract (T-OCC) and increased FD in the left AF. These studies have provided preliminary evidence of FBA-informed WM alterations in mood disorders, highlighting the potential for further research using this analysis framework.

The present study is the first to compare FBA-derived WM micro- and macrostructural properties in BD-II with those in UD and HC. Importantly, datasets from the two existing studies comparing BD-II and UD (Mak et al., 2021; Manelis et al., 2021) were combined and harmonized to generate a larger sample. Relationships between WM alterations and clinical characteristics were also examined. Sensitivity analyses in unmedicated participants were conducted to address the potential impact of psychotropic medications on WM changes (Benedetti et al., 2011; Favre et al., 2019; Mamah et al., 2019). We hypothesized that (1) BD-II and UD would show decreased FD, FC, and FDC compared to HC, (2) BD-II would display more pronounced WM changes than UD, (3) in BD-II and UD, FBA metrics would correlate with mood symptoms, number of mood episodes, and the age of illness onset, (4) these effects would be most pronounced in major WM tracts such as IFO, ILF, SLF, CG, and CC, and, (5) compared to the main results, the sensitivity analyses in the unmedicated subset would show different patterns of WM disruptions, reflecting the effect of psychotropic medications on WM integrity.

## 2. Methods

### 2.1. Participants

#### Original datasets

The original datasets were obtained from the study by Manelis et al. (2021) at the University of Pittsburgh, approved by the University of Pittsburgh (IRB number: STUDY19090330) and Carnegie Mellon University (IRB number: CR00000952) Institutional Review Boards, and the study by Mak et al. (2021) at the Chinese University of Hong Kong (CUHK), approved by the New Territories East Cluster – CUHK Clinical Research Ethics Committee (CREC number: 2014.168). The Pittsburgh dataset included 65 participants, comprised of 18 individuals with bipolar II disorder (BD-II), 23 with unipolar depression (UD), and 24 healthy controls (HC). The Hong Kong (HK) dataset included 81 participants, with 27 each of BD-II, UD, and HC. Details of recruitment and inclusion/exclusion criteria have been reported in previous publications (Mak et al., 2021; Manelis et al., 2021) and presented in supplementary material S1. All eligible participants provided informed consent.

#### Combined dataset

The datasets were combined and analysed according to the data collaboration agreement between CUHK and the University of Pittsburgh. Twelve participants were excluded due to rapid cycling (n=11) or imaging artifacts (n=1). The combined dataset included 134 participants (Pittsburgh=64; HK=70), comprised of 33 BD-II, 50 UD, and 51 HC. For the sensitivity analyses, 25 individuals (BD-II=14; UD=11) currently using psychotropic medications were excluded, yielding a subset of 109 unmedicated participants (Pittsburgh=39; HK=70) with 19 BD-II, 39 UD, and 51 HC.

### 2.2. Clinical assessments

#### Pittsburgh

Diagnostic assessments were conducted by trained interviewers, supervised by a psychiatrist, according to DSM-5 criteria using the Mini International Neuropsychiatric Interview (MINI) 7.0 (Lecrubier et al., 1997). Age of onset, number of depressive and hypomanic episodes, family history of BD, and medication history were acquired by self-report. Past-week affective symptoms were assessed using the 25-item Hamilton Rating Scale for Depression (HRSD-25) (Miller et al., 1985) and Young Mania Rating Scale (YMRS) (Young et al., 1978) on the day of scan. Premorbid intelligence was estimated with the National Adult Reading Test (NART) (Nelson & Willison, 1991).

#### HK

Diagnostic assessments were conducted by trained interviewers, supervised by a psychiatrist, using the Chinese bilingual version of the Structured Clinical Interview for DSM-IV Axis I Disorders (SCID-I) (Benazzi & Akiskal, 2003; Mak, 2009; So et al., 2003). Lifetime affective episodes were enquired on a year-by-year basis. Age of onset and family history of BD were acquired by self-report. Medication history was enquired from participants and caregivers, confirmed with public hospital registry. Past-week affective symptoms were rated using the interviewer-administered Montgomery-Åsberg Depression Rating Scale (MADRS) (Montgomery & Åsberg, 1979) and YMRS (Young et al., 1978). General intelligence was measured with the three-subtest short-form Wechsler Adult Intelligence Scale-III (WAIS-III) (Chan et al., 2005).

### 2.3. Harmonization of clinical data

#### Depressive symptoms

The 17-item HRSD score was derived from HRSD-25 by selecting relevant items (Hamilton, 1986), which was converted to the MADRS scale using a published conversion scheme (Carmody et al., 2006). The harmonized score, referred to as “depressive symptoms”, ranged from 0 to 60.

#### Intellectual ability

IQ score was z-transformed using the population mean and standard deviation (SD). NART IQ had population mean of 100 and SD of 15. Since the standardization manual for the three-subtest short-form WAIS-III was unavailable, we approximated the population statistics from the results of Chan et al. (2005), yielding mean of 33.1 and SD of 5.1. To avoid confusion with the official IQ score, the standardized score is referred to as “IQ z-score”.

### 2.4. Image acquisition

#### Pittsburgh

Diffusion-weighted images (DWI) were collected using a Siemens Verio 3T scanner with a 32-channel head coil, using a multi-band (MB) sequence (TR=3033ms, TE=124.6ms, FOV=220×220mm^2^, 68 axial slices, resolution=2×2×2mm^3^, flip angle=90°, acceleration factor=4). Diffusion gradients were applied along 150 directions with b=2000s/mm^2^, with 16 non-diffusion-weighted (b=0s/mm^2^) images.

#### HK

Imaging data were collected using a Philips Achieva 3.0T TX scanner with an 8-channel head coil. Structural images were acquired using a T1-weighted sequence (TR=7.4ms, TE=3.4ms, FOV=250×250mm^2^, 285 sagittal slices, slice thickness=0.6mm, acquisition matrix=240×240, flip angle=8°). DWI were acquired using a single shot echo planar imaging (EPI) sequence (TR=8912ms, TE=60ms, FOV=224×224mm^2^, 70 axial slices, resolution=2×2×2mm^3^, flip angle=90°, acceleration factor=2.5). Diffusion gradients were applied along 32 directions with b=1000s/mm^2^, with an acquisition without diffusion weighting (b=0s/mm^2^).

### 2.5. Data preprocessing

DWI were preprocessed using MRtrix 3.0.4 and FSL (Jenkinson et al., 2012; Tournier et al., 2019). Images were denoised using *dwidenoise* and Gibbs ringing artifacts removed using *mrdegibbs* (Kellner et al., 2016; Veraart et al., 2016). For the Pittsburgh dataset, susceptibility-induced distortions, eddy currents, and motion artifacts were corrected using *dwifslpreproc*, leveraging the *topup* and *eddy* commands of FSL (Andersson & Sotiropoulos, 2016; Smith et al., 2004). For the HK dataset with single phase encoding, the denoised, unringed b=0 data and the T1-weighted image were used to synthesize an “undistorted” b=0 image using Synb0-DISCO (Schilling et al., 2020; Schilling et al., 2019), which was utilized to improve distortion correction using *dwifslpreproc* (Andersson & Sotiropoulos, 2016; Smith et al., 2004). For both datasets, bias field was corrected using *dwibiascorrect* (Zhang et al., 2001).

### 2.6. Fixel-based analysis (FBA)

FBA was performed using MRtrix3 3.0.4, according to the MRtrix3 documentation and B.A.T.M.A.N. manual (Tahedl et al., 2025; Tournier et al., 2019). Response functions (RF) of white matter (WM), grey matter (GM), and cerebrospinal fluid (CSF) were estimated using *dwi2response* (Dhollander et al., 2019; Dhollander et al., 2016). Group-level RF was calculated by averaging participants’ RFs *within* each dataset to preserve site-specific scaling factors. Preprocessed DWI were upsampled to 1.25×1.25×1.25mm^3^ for better resolution in downstream processes. Brain masks were generated using the FSL *bet* command (Bastiani et al., 2019).

The single-shell 3-tissue constrained spherical deconvolution (SS3T-CSD) algorithm was used for FOD estimation in all three tissue types (Dhollander & Connelly, 2016). While relying on single-shell data, SS3T-CSD outperformed the multi-shell multi-tissue CSD (MSMT-CSD) in estimating FODs within abnormal WM structures (Aerts et al., 2019). The *ss3t_csd_beta1* command of the MRtrix3Tissue toolbox was applied for each dataset with the site-specific average RF (Dhollander & Connelly, 2016). Bias field correction and global intensity normalization were performed jointly using *mtnormalise*. WM FOD template was created with a subset of 30 participants (10 per group) using *population_template*. The template mask was generated by registering all participants’ masks to the template space and calculating their intersection to prevent inclusion of non-brain voxels in any participant. Each participant’s fixel image was derived from the FOD (transformed to template space) using *fod2fixel* (Smith et al., 2013). Fixels were reoriented using *fixelreorient* based on the native-to-template transformation matrix. Template fixel mask was generated from the FOD template using *fod2fixel*, utilized for assigning fixel correspondence across participants using *fixelcorrespondence* (Smith et al., 2013).

Three metrics were derived for statistical testing. FD was computed during fixel estimation as the FOD integral of the corresponding fixel (Raffelt et al., 2012). FC, defined as the morphological change perpendicular to the fibre (fixel) orientation, was derived by applying *warp2metric* on the native-to-template warps, and were log-transformed to centre around zero (Raffelt et al., 2017). FDC was calculated as the product of FD and FC. To account for between-site heterogeneity, diffusion metrics were harmonized using ComBat, which effectively removes inter-site technical variability while preserving biological differences (Orlhac et al., 2022).

### 2.7. Tract segmentation and connectivity-based fixel smoothing

TractSeg was used to obtain streamlines in 72 well-known WM tracts (Wasserthal et al., 2018). Peak values of the FOD template were extracted using *sh2peaks* of MRtrix3 (Jeurissen et al., 2013), and passed to the *TractSeg* command for tract segmentation, tract endings definition, and creation of study-specific tract orientation maps. High-resolution bundle-specific tractography was performed using the *Tracking* command, creating 10000 fibres per tract, which were also used to estimate fixel-fixel connectivity to guide data smoothing with a 10-mm Gaussian filter using *fixelfilter*.

### 2.8. Statistical analyses

#### Demographic and clinical data

Group differences and comparisons between medicated and unmedicated individuals were analysed using ANOVA, t-test, or chi-square test, whichever appropriate, implemented in R 4.2.3 (R Core Team, 2013). Post-hoc t-tests were corrected for the false discovery rate (FDR). 2(group)x 2(medication) ANOVA were also conducted to examine the effects of diagnostic group and medication status, and their interaction, on clinical variables in BD-II and UD. Results are presented in supplementary material S3.

#### FBA

Non-parametric permutation testing was used to assess group differences in FD, FC, and FDC per tract, using *fixelcfestats* of MRtrix3 with 3000 permutations (Raffelt et al., 2015). Three pairwise comparisons between BD-II, UD and HC were tested. Age, sex, and IQ z-score were demeaned and included as covariates. A variance block of site was supplied to control for residual variances. The fixel-fixel connectivity matrix was used for connectivity-based fixel enhancement (CFE) to identify clustered statistics without an arbitrary threshold (Raffelt et al., 2015; Smith et al., 2019). Familywise error rate (FWER) was used for fixel-wise correction. The resulting statistical maps were filtered with Bonferroni-corrected p<0.017 to account for three tests. Group effects were represented by the maximum *t* and minimum *p* per tract.

#### FBA-clinical correlation

For each tract with significant group differences, the average FD/FC/FDC across significant fixels was computed for each participant of the corresponding diagnostic group using *mrstats*, and the partial correlations with clinical variables, including depressive symptoms, YMRS, number of MDE, and illness duration, were analysed using *pcor.test* in R, controlling for age, sex, and IQ z-score, with FDR correction for multiple testing (Kim, 2015).

#### Sensitivity analyses in the unmedicated subset

Two sets of sensitivity analysis were performed in the unmedicated subset (n=109) to examine whether the observed findings were caused by medication effects. First, for each tract with significant group differences, the average metric values across significant fixels were computed for all unmedicated participants using *mrstats*, which were compared between the corresponding groups using t-test, implemented in R. Second, non-parametric permutation tests of group differences in FBA metrics within the unmedicated subset were performed using *fixelcfestats* according to the procedures described above.

## 3. Results

### 3.1. Demographic and clinical characteristics

#### Full dataset

No significant group differences were observed in age, sex, and IQ z-score. BD-II had higher current depressive (harmonized score; *p*=0.048) and manic symptom scores (YMRS; *p*=0.001) than UD. BD-II participants reported earlier illness onset (*p*=0.02) than UD. Family history of BD was more common in BD-II than in UD (*p*=0.001). There were no significant differences in the number of major depressive episodes (MDE) lifetime and medication status between BD-II and UD. Results are summarized in Table 1. Supplementary material S2 summarizes site-specific statistics.

**Table 1.** Demographic and clinical characteristics

| Variable | Mean (SD) / Count (%) |  |  |  |  | Test | Group differences |  |  |  |
| --- | --- | --- | --- | --- | --- | --- | --- | --- | --- | --- |
|  | Full dataset |  | Unmedicated subset |  | HC<br>(n = 51) |  | Full dataset |  | Unmedicated subset |  |
|  | BD-II<br>(n = 33) | UD<br>(n = 50) | BD-II<br>(n = 19) | UD<br>(n = 39) |  |  | F / t / X<sup>2</sup> (df) | p | F / t / X<sup>2</sup> (df) | p |
| Age | 24.1 (4.2) | 24.8 (5.6) | 24.3 (4.5) | 24.9 (5.4) | 24.5 (5.5) | ANOVA | 0.2 (2,128) | 0.81 | 0.1 (2,103) | 0.89 |
| Sex |  |  |  |  |  | Chi-square | 1.3 (2) | 0.52 | 2.6 (2) | 0.28 |
| Male | 8 (24.2%) | 14 (28.0%) | 3 (15.8%) | 11 (28.2%) | 18 (35.3%) |  |  |  |  |  |
| Female | 25 (75.8%) | 36 (72.0%) | 16 (84.2%) | 28 (71.8%) | 33 (64.7%) |  |  |  |  |  |
| IQ z-score | 0.3 (1.0) | 0.0 (1.4) | 0.0 (1.2) | 0.0 (1.5) | 0.5 (1.1) | ANOVA | 3.0 (2,128) | 0.05 | 4.2* (2,103) | 0.02 |
|  |  |  |  |  |  | T-test (BD-II–HC) | -1.1 (75.5) | 0.28 | -1.6 (31.0) | 0.18 |
|  |  |  |  |  |  | T-test (UD–HC) | -2.2 (92.9) | 0.10 | -2.6* (66.7) | 0.03 |
|  |  |  |  |  |  | T-test (BD-II–UD) | 1.1 (80.9) | 0.28 | 0.7 (45.6) | 0.49 |
| Depressive symptoms | 27.4 (8.6) | 24.1 (5.1) | 26.4 (9.6) | 25.0 (5.0) | 0.2 (1.1) | T-test<sup>^</sup> | 2.0* (47.0) | 0.048 | 0.6 (22.9) | 0.56 |
| YMRS | 4.1 (3.8) | 1.6 (1.8) | 4.7 (4.4) | 1.6 (1.9) | 0.2 (0.6) | T-test<sup>^</sup> | 3.5** (41.8) | 0.001 | 3.0** (21.4) | 0.007 |
| Psychotropic drug use |  |  |  |  |  | Chi-square<sup>^</sup> | 3.0 (1) | 0.08 | / | / |
| Absent | 19 (57.6%) | 39 (78.0%) | / | / | 51 (100%) |  |  |  | / | / |
| Present | 14 (42.4%) | 11 (22.0%) | / | / | 0 (0%) |  |  |  | / | / |
| Family history of BD |  |  |  |  |  | Chi-square<sup>^</sup> | 10.8** (1) | 0.001 | 9.8** (1) | 0.002 |
| Absent | 22 (66.7%) | 48 (96.0%) | 11 (57.9%) | 37 (94.9%) | 51 (100%) |  |  |  |  |  |
| Present | 11 (33.3%) | 2 (4.0%) | 8 (42.1%) | 2 (5.1%) | 0 (0%) |  |  |  |  |  |
| Lifetime MDE | 3.2 (1.9) | 3.6 (3.9) | 2.3 (1.1) | 3.3 (4.1) | / | T-test<sup>^</sup> | -0.5 (76.2) | 0.62 | -1.4 (48.0) | 0.17 |
| Lifetime HME | 2.4 (1.7) | / | 2.2 (1.7) | / | / | N/A | / | / | / | / |
| Age of illness onset | 17.8 (4.0) | 20.4 (6.2) | 18.3 (4.2) | 20.8 (6.0) | / | T-test<sup>^</sup> | -2.3* (81.0) | 0.02 | -1.8 (48.5) | 0.07 |
| Illness duration (year) | 7.2 (4.0) | 5.4 (4.3) | 6.9 (4.8) | 5.1 (4.7) | / | T-test<sup>^</sup> | 2.0<sup>†</sup> (71.7) | 0.05 | 1.4 (35.2) | 0.18 |

BD-II, bipolar-II depressed; UD, unipolar depressed; HC, healthy control; YMRS, Young Mania Rating Scale; BD, bipolar disorder; MDE, major depressive episode; HME, hypomanic episode
Statistically significant at $p < (*)$ 0.05 and $(**)$ 0.01 levels
<sup>^</sup>Comparison between BD-II and UD

#### Unmedicated subset

No significant group differences were observed in age and sex. BD-II did not differ significantly from UD and HC in IQ z-score, whereas UD showed significantly lower IQ z-score than HC (*p*=0.03). BD-II had higher YMRS than UD (*p*=0.007). No significant differences were found in current depressive symptoms, number of MDE, age of illness onset, and illness duration between BD-II and UD. Family history of BD was more prevalent in BD-II than UD (*p*=0.002). Results are summarized in Table 1.

#### Comparison between medicated and unmedicated individuals

Please refer to supplementary materials S3 for the results of two-way ANOVA examining the interaction effects of group and medicated status on the clinical variables. Overall, medicated individuals had significantly higher IQ z-score (*p*<0.001) and more lifetime MDE (*p*=0.02) compared to unmedicated individuals. Within UD, medicated individuals showed significantly higher IQ z-score (*p*=0.001) and lower depressive symptom score (*p*=0.005) compared to unmedicated individuals. Within BD-II, medicated individuals had more lifetime MDE than unmedicated individuals (*p*=0.002). Results are summarized in Table 2.

**Table 2.** Comparison of sample characteristics between medicated and unmedicated individuals

| Variable | Overall |  |  |  | BD-II |  |  |  | UD |  |  |  |
| --- | --- | --- | --- | --- | --- | --- | --- | --- | --- | --- | --- | --- |
|  | Mean (SD) / Count (%) |  | Comparison |  | Mean (SD) / Count (%) |  | Comparison |  | Mean (SD) / Count (%) |  | Comparison |  |
| | Medicated | Unmedicated | $t / X^2$ (df) | $p$ | Medicated | Unmedicated | $t / X^2$ (df) | $p$ | Medicated | Unmedicated | $t / X^2$ (df) | $p$ |
| Age | 24.1 (5.3) | 24.7 (24.7) | 0.5 (43.7) | 0.62 | 23.8 (3.9) | 24.3 (4.5) | 0.3 (30.0) | 0.75 | 24.5 (6.8) | 24.9 (5.4) | 0.2 (13.6) | 0.84 |
| Sex | - | - | 0.2 (1) | 0.64 | - | - | 0.8 (1.0) | 0.36 | - | - | < 0.001 (1) | > 0.99 |
| *Male* | 8.0 (0.3) | 14.0 (14.0) | - | - | 5.0 (0.4) | 3.0 (0.2) | - | - | 3.0 (0.3) | 11.0 (0.3) | - | - |
| *Female* | 17.0 (0.7) | 44.0 (44.0) | - | - | 9.0 (0.6) | 16.0 (0.8) | - | - | 8.0 (0.7) | 28.0 (0.7) | - | - |
| IQ (z-score) | 0.6 (0.4) | -0.2 (-0.2) | -4.0*** (73.5) | < 0.001 | 0.6 (0.4) | 0.0 (1.2) | -2.1 (23.1) | 0.05 | 0.7 (0.4) | -0.3 (1.5) | -3.4** (48.0) | 0.001 |
| Depressive symptoms | 25.2 (7.0) | 25.5 (25.5) | 0.15 (44.4) | 0.88 | 28.8 (7.0) | 26.4 (9.6) | -0.8 (31.0) | 0.41 | 20.7 (3.8) | 25.0 (5.0) | 3.1** (20.9) | 0.005 |
| YMRS | 2.3 (2.4) | 2.7 (2.7) | 0.5 (59.8) | 0.61 | 3.1 (2.7) | 4.7 (4.4) | 1.3 (30.4) | 0.21 | 1.3 (1.6) | 1.6 (1.9) | 0.7 (19.3) | 0.52 |
| Family history of BD | - | - | 0.1 (1) | 0.78 | - | - | 0.8 (1.0) | 0.38 | - | - | < 0.001 (1) | > 0.99 |
| *Absent* | 22.0 (0.9) | 48.0 (48.0) | - | - | 11.0 (0.8) | 11.0 (0.6) | - | - | 11.0 (1.0) | 37.0 (0.9) | - | - |
| *Present* | 3.0 (0.1) | 10.0 (10.0) | - | - | 3.0 (0.2) | 8.0 (0.4) | - | - | 0.0 (0.0) | 2.0 (0.1) | - | - |
| Lifetime MDE | 4.5 (2.4) | 3.0 (3.0) | -2.3* (63.1) | 0.02 | 4.5 (2.1) | 2.3 (1.1) | -3.5** (18.3) | 0.002 | 4.5 (2.9) | 3.3 (4.1) | -1.2 (22.4) | 0.26 |
| Lifetime HME | - | - | - | - | 2.8 (1.8) | 2.2 (1.7) | -1.0 (27.3) | 0.31 | - | - | - | - |
| Age of illness onset | 18.0 (5.4) | 20.0 (20.0) | 1.5 (46.4) | 0.13 | 17.1 (3.7) | 18.3 (4.2) | 0.8 (29.9) | 0.40 | 19.0 (7.1) | 20.8 (6.0) | 0.8 (14.2) | 0.46 |
| Illness duration (year) | 7.1 (2.3) | 5.7 (5.7) | -1.8 (80.1) | 0.08 | 7.6 (2.6) | 6.9 (4.8) | -0.5 (29.0) | 0.60 | 6.5 (1.6) | 5.1 (4.7) | -1.5 (45.9) | 0.15 |

BD-II, bipolar-II depressed; UD, unipolar depressed; YMRS, Young Mania Rating Scale; BD, bipolar disorder; MDE, major depressive episode; HME, hypomanic episode
a. Harmonized scale
Statistically significant at $p < (*)$ 0.05, $(**)$ 0.01, and $(***)$ 0.001 levels

### 3.2. Fixel-based analysis (FBA)

The level of significance was adjusted to 0.017 (0.05/3) for three between-group comparisons using Bonferroni correction. Significant findings are summarized in Table 3.

**Table 3.**
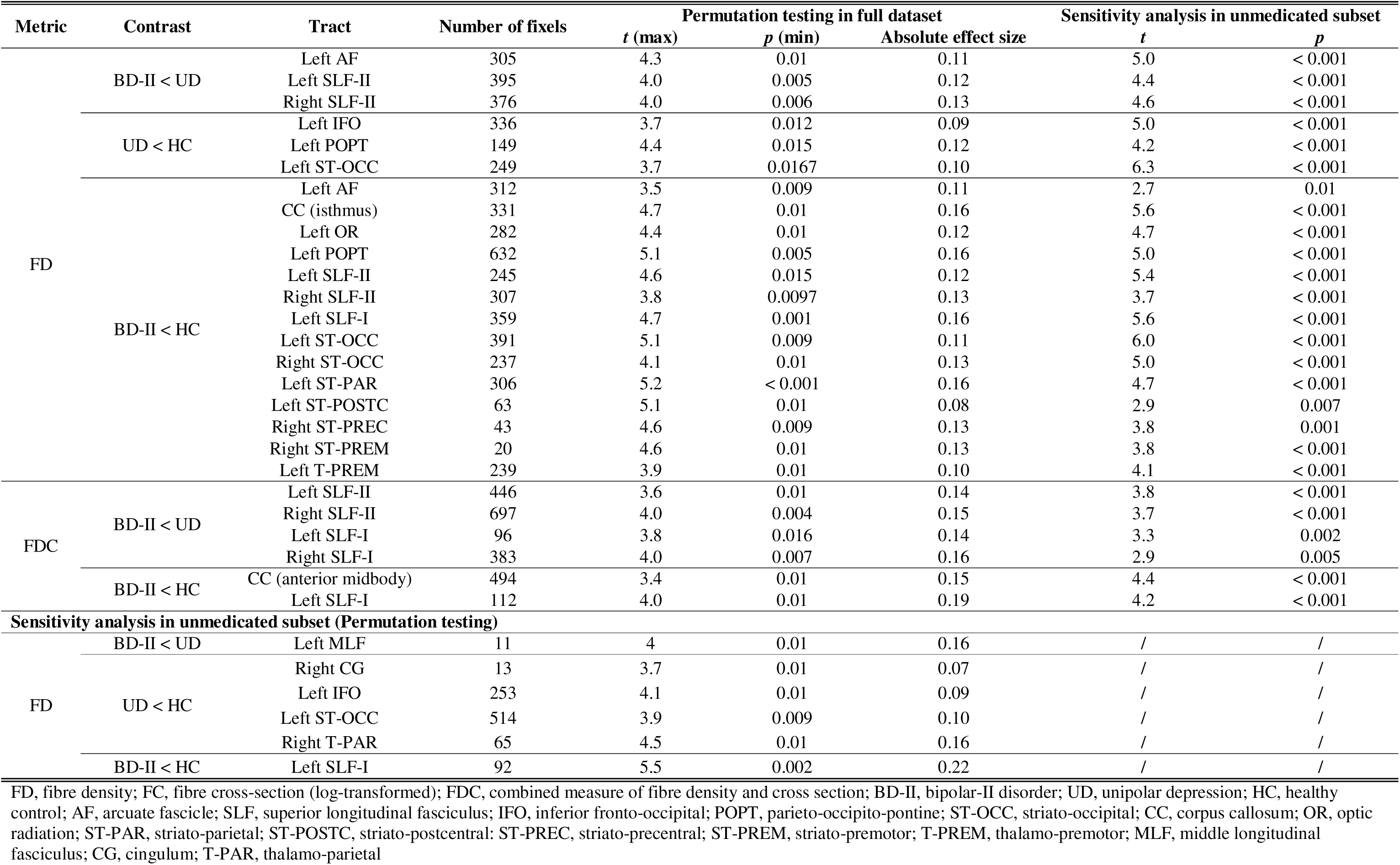
Results of fixel-based analysis and sensitivity analyses in unmedicated subset

### Full dataset

#### FD

BD-II showed significantly lower FD compared to UD in the left AF and bilateral SLF-II. BD-II showed reduced FD compared to HC in 14 tracts, including the left arcuate fascicle (AF), left superior longitudinal fasciculus I (SLF-I), bilateral SLF-II, left optic radiation (OR), left parietal-occipital-pontine tract (POPT), left thalamo-premotor tract (T-PREM), six striato-cortical tracts, and the isthmus of CC. UD displayed reduced FD relative to HC in the left inferior fronto-occipital tract (IFO), left POPT, and left striato-occipital tract (ST-OCC; marginally significant).

#### FC

There were no significant differences of FC in any tracts.

#### FDC

BD-II exhibited lower FDC compared to UD in bilateral SLF-I and II, and compared to HC in the left SLF-I and the anterior midbody of CC. There was no significant difference in FDC between UD and HC.

### Sensitivity analyses in the unmedicated subset

#### T-tests

Results of t-tests indicated that, for all significant FBA findings in the full dataset, the group differences in FD/FDC remained significant in the unmedicated subset (*ps*<0.05) (Table 3).

#### Permutation testing (fixelcfestats)

In the unmedicated subset, BD-II showed significantly lower FD in the left MLF compared to UD and in the left SLF-I compared to HC. UD exhibited reduced FD compared to HC in the left IFO, right cingulum (CG), left ST-OCC, and right thalamo-parietal tract (T-PAR). No significant differences of FC and FDC were observed in any tracts.

### 3.3. FBA-clinical correlations

#### Full dataset

In BD-II, illness duration negatively correlated with FD in the left AF, left POPT, and right ST-OCC, whereas lifetime number of MDE positively correlated with FDC in the left SLF-I (FDR-corrected *ps*<0.05). No significant correlation was found in UD. Figure 1 illustrates the group differences and correlations in these tracts.

#### Unmedicated subset

There was no significant FBA-clinical correlation in the unmedicated individuals.

## 4. Discussion

We have reported on the first study to use fixel-based analysis (FBA) to investigate white matter (WM) abnormalities in individuals with bipolar II disorder (BD-II) compared to those with unipolar depression (UD) and healthy controls (HC), using a combined and harmonized dataset from two studies (Mak et al., 2021; Manelis et al., 2021). We found more extensive WM disruptions associated with clinical characteristics (illness duration and the number of depressive episodes) in BD-II compared to UD and HC. This is a valuable contribution to the relatively sparse WM literature on BD-II plagued by inconsistent findings with small sample sizes owing to recruitment challenges, assessment difficulties, and heterogeneity in clinical and demographic characteristics (Ambrosi et al., 2016; Ambrosi et al., 2013; Caseras et al., 2015; Foley et al., 2018; Ha et al., 2009; Kurumaji et al., 2017; Liu et al., 2010; Mak et al., 2021; Thiel et al., 2024; Yip et al., 2013).

We observed shared FD reductions in the left POPT and left ST-OCC in BD-II and UD compared to HC. Decreased FD in the left ST-OCC is consistent with previous FBA findings in UD and mixed BD subtypes (Manelis et al., 2025). While disruptions of the POPT have not been reported in DTI and FBA studies of UD, DTI studies have shown reduced FA in BD (Cao et al., 2025). ST-OCC integrates visual and semantic processing to support verbal fluency (Egorova-Brumley et al., 2022). POPT connects the parietal and occipital lobes with the pontine nuclei, integrating sensory information to support motor coordination (Calixto et al., 2025; Rousseau et al., 2022). These findings suggest shared WM microstructural disruptions in BD-II and UD that may underly the shared impairments in visual, semantic, and motor processing and integration commonly observed in mood disorders (Bubl et al., 2010; Henry & Crawford, 2005; O’Bryan et al., 2014; Raucher-Chéné et al., 2017).

BD-II displayed extensive FD decreases in other major WM tracts and decreased FDC in bilateral SLF-I and II and the anterior midbody of the CC. FD reductions in the right SLF-II and left optic radiation (OR) are consistent with previous FBA findings in BD (Manelis et al., 2025). Decreased FD and FDC in the SLF and CC also broadly align with DTI findings of decreased FA within these tracts in BD (Ambrosi et al., 2013; Maller et al., 2014; Yip et al., 2013). To our knowledge, there are no prior reports of WM disruptions in the left AF, left thalamo-premotor tract (T-PREM), and multiple striato-cortical tracts in BD-II. Similar to the present analysis, Manelis et al. (2025) showed decreased FD in the SLF-II in BD vs UD, but their findings in the left AF was opposite to the current findings. This may reflect the differences of bipolar subtypes or current mood states of the participants in the present study compared to those in Manelis et al. (2025). Notably, we present the first evidence of FDC reductions in BD-II and, more broadly, in BD, indicating not only within-voxel fibre degeneration but also tract-level disruptions.

The SLF-I connects the superior parietal lobe and precuneus to the superior frontal gyrus and supplementary motor area, whereas the SLF-II extends from the inferior parietal lobe to the middle frontal gyrus. Both support visuospatial attention through integrating visual processing with motor planning (Vergani et al., 2021; Wang et al., 2016). The OR, extending from the lateral geniculate to the primary visual cortex, is implicated in visual information processing and working memory (Wang et al., 2024). The AF connects frontal, parietal, and temporal cortices, including the Broca’s and Wernicke’s areas, which supports language processing (Fernández-Miranda et al., 2015; Ivanova et al., 2021). The isthmus of CC connects parietal and occipital regions of both hemispheres, supporting sensorimotor integration (Güntürkün et al., 2020; Maharajh et al., 2024). The anterior midbody of CC connects interhemispheric prefrontal and motor cortices, implicated in gesture imitation and moral reasoning in which damage would hinder self-other relation and impair social interaction (Chao et al., 2009; Fabri & Polonara, 2023; Miller et al., 2010; Pierpaoli et al., 2018). T-PREM is involved in motor information relay that supports motor control, whereas the striato-cortical pathways are implicated in the motor control of speech and serial-order working memory (Wang et al., 2021; Watkins & Jenkinson, 2016; Ye et al., 2020). These widespread disruptions in WM tracts may underlie the impairments in visuomotor and language processing linked to several components of executive dysfunction such as spatial attention, working memory, and verbal fluency in BD (Hsiao et al., 2009; Raucher-Chéné et al., 2017; Torrent et al., 2006; Xu et al., 2012).

Participants with UD exhibited reduced FD in the left IFO, which broadly aligns with DTI findings of FA reduction (Liao et al., 2013). IFO connects the parietal and occipital lobes to the frontal lobe via the insula, which subserves the processing of emotion in faces (Conner et al., 2018; Martino et al., 2010; Philippi et al., 2009). Damage in this tract may explain deficits in facial emotion recognition in UD (Dalili et al., 2015). Our findings suggest that axonal damage in UD primarily affects the visual processing pathways responsible for comprehending visual information and generating responses, potentially underscoring emotion-related perceptual, cognitive, and decision biases in depression (Bubl et al., 2010; Dalili et al., 2015; Kohler et al., 2011). Contrary to Lyon et al. (2019), we found no significant FD reductions in the CC in UD. This may result from differences in demographics and clinical profiles, as our UD group had a younger mean age, shorter illness duration, and fewer depressive episodes.

In BD-II, illness duration negatively correlated with FD in the left AF, left POPT, and right ST-OCC, suggesting progressive WM disruptions with illness chronicity. This aligns with the neuroprogression model of bipolar disorders which posits progressive clinical deterioration occurring along with structural and functional brain changes in the context of a host of accumulated metabolic, immune and neurotrophic aberrations (Egorova-Brumley et al., 2022; Ivanova et al., 2021; Rousseau et al., 2022). Unexpectedly, we observed *positive* correlation between depression recurrence and FDC in the left SLF-I. This may indicate adaptive WM reorganization, potentially through utilization of spatial attention to compensate for impaired sensory processing. Nevertheless, factors such as cross-sectional design, multifaceted nature of FDC, and potential confounding effects (e.g., illness severity and medications) may have influenced the findings, emphasizing the need for longitudinal research in larger samples to clarify the nature and trajectories of these changes.

Similar to our earlier report on the HK sample (Mak et al., 2021), the present analysis did not yield significant correlations between lifetime number of hypomanic episodes and WM abnormalities. This contrasts with mania, where patients with a greater number of episodes show more extensive WM changes (Thiel et al., 2025). However, it is premature to conclude that recurrence of hypomania is unrelated to WM alterations, given its inherently transient nature, which likely requires a longer illness course and larger sample size to detect significant associations. Future prospective longitudinal imaging studies with frequent sampling of affective measures should help resolve current uncertainties regarding the chronology of hypomania.

Our respective recruitment strategies allowed to perform sensitivity analyses in an unmedicated subset to address potential effects of psychotropic medications on WM changes. First, group differences within significant fixels in the main FBA remained significant in the unmedicated individuals. Second, in the permutation analysis with unmedicated individuals only, BD-II exhibited reduced FD in the left SLF-I compared to HC and in the left middle longitudinal fasciculus (MLF) compared to UD, whereas unmedicated UD showed reduced FD in the left IFO, left ST-OCC, right cingulum (CG), and right thalamo-parietal tract (T-PAR) relative to HC. FD reductions in BD-II were less extensive in unmedicated individuals, consistent with previous findings of less pronounced WM disruptions in unmedicated individuals who are often less symptomatic than those receiving psychotropic medications (Manelis et al., 2025; Versace et al., 2008), although there have also been reports of protective medication effects on WM integrity (Benedetti et al., 2011; Favre et al., 2019). Moreover, the loci of WM alterations partially differed from those observed in the full dataset. Unmedicated individuals with BD-II showed reduced FD in the left MLF, which connects the superior temporal gyrus with the parietal and occipital cortices, supporting verbal-auditory learning (Latini et al., 2021). In unmedicated individuals with UD, decreased FD was observed in the right CG, which is a prominent cortico-limbic tract implicated in emotion regulation (Bubb et al., 2018). Nevertheless, the small fixel count and sample size warrant caution in interpretation. Furthermore, it is premature to conclude that the differing results are treatment-related, as unmedicated individuals also had fewer depressive episodes (BD-II) and more severe depressive symptoms (UD) that may also account for the observed differences. Unfortunately, the small sample size, uneven distribution of medicated individuals across sites, and heterogeneous medication types and dosages hinder direct comparison of WM integrity by medication status. Still, our findings provide clear evidence of WM alterations in unmedicated individuals.

To date, most studies in bipolar disorders (BD) focused on individuals with BD-I or mixed BD-I/BD-II subtypes. Few investigated WM abnormalities in BD-II, largely due to difficulty in recruiting and retaining participants. Some studies showed decreased FA in BD-II relative to HC (Ambrosi et al., 2016; Ambrosi et al., 2013; Ha et al., 2009; Kurumaji et al., 2017; Liu et al., 2010; Thiel et al., 2024; Yip et al., 2013), while others reported no significant WM alterations (Caseras et al., 2015; Foley et al., 2018; Mak et al., 2021). Such discrepancies may result from the wide range of illness durations (6.5 – 23.9 years), heterogeneous medication status, and differing imaging parameters and analysis approaches. The present study provides robust evidence of WM alterations in BD-II compared to UD and HC, utilizing a harmonized dataset with a young sample and short illness duration as well as rigorous FBA technique, while also addressing potential medication effects.

Nevertheless, there are several limitations. First, a prospectively and jointly planned data collection protocol would be more rigorous than the current joint data analysis and harmonization protocol, despite broad similarities in our clinical and imaging data acquisition designs. It behoves upon the global bipolar research community to join in collaborative efforts to better understand bipolar II disorder, given the specific recruitment challenges associated with participant identification compared to other affective conditions. Second, the cross-sectional design hinders investigation of causal inferences between illness phenomenology and WM pathology. Third, while excluding participants with rapid cycling increases sample homogeneity, it also under-represents the broader bipolar spectrum. Fourth, the Pittsburgh and HK studies assessed premorbid and current intellectual abilities, respectively. This likely introduced harmonization challenges, and the current IQ measure may also be confounded by WM disruptions. Future studies should use standardized IQ measures, preferably premorbid IQ. Lastly, future work should acquire multilshell data that not only improve FOD estimation for better FBA accuracy (Dhollander et al., 2019) but also support other advanced analytical approaches such as NODDI.

## 5. Conclusions

This is the first FBA study to investigate white matter (WM) alterations in bipolar II disorder (BD-II) and unipolar depression (UD) compared to HC. Shared mechanisms involve pathways implicated in visuomotor integration and spatial attention, including the left parieto-occipito-pontine tract (POPT) and striato-occipital tract (ST-OCC). While WM abnormalities in UD were observed in visuomotor and visuospatial attention pathways that may underscore perceptual and cognitive biases, BD-II demonstrates more widespread disruptions across association, projection, and commissural tracts supporting sensory, motor, semantic, and executive processing. Importantly, the extensive WM alterations observed early in the illness course highlight the need for early identification and intervention. Albeit less pronounced, significant differences in FD reductions in BD-II and UD were also observed in an unmedicated subset. Correlation analyses further suggest potential degenerative and compensatory mechanisms specific to BD-II related to overall illness burden, but not hypomanic symptomatology, warranting prospective, well-sampled longitudinal studies to track symptom trajectories alongside WM alterations across different mood states.

## Author contributions

**Idy W. Y. Chou:** Formal analysis, Methodology, Software, Validation, Visualization, Writing – original draft, Writing – review & editing

**Anna Manelis:** Conceptualization, Data curation, Funding acquisition, Investigation, Methodology, Project administration, Supervision, Writing – review & editing

**Holly A. Swartz:** Conceptualization, Supervision, Validation, Writing – review & editing

**Owen N. W. Leung:** Writing – review & editing

**Mary L. Phillips:** Conceptualization, Supervision, Writing – review & editing

**Suzanne H. W. So:** Data curation, Investigation, Methodology, Writing – review & editing

**Winnie C. W. Chu:** Data curation, Investigation, Methodology, Writing – review & editing

**Hanna Lu:** Project administration, Resources, Supervision, Writing – review & editing

**Linda C. W. Lam:** Data curation, Methodology, Project administration, Resources, Supervision, Writing – review & editing

**Arthur D. P. Mak:** Conceptualization, Data curation, Funding acquisition, Investigation, Methodology, Project administration, Supervision, Writing – review & editing

All authors have read and approved the final version of the manuscript and agreed to be accountable for all aspects of this work.

## Data Availability

All data produced in the present study are available upon reasonable request to the authors.

## Acknowledgements

The authors thank participants for taking part in this research study.

---

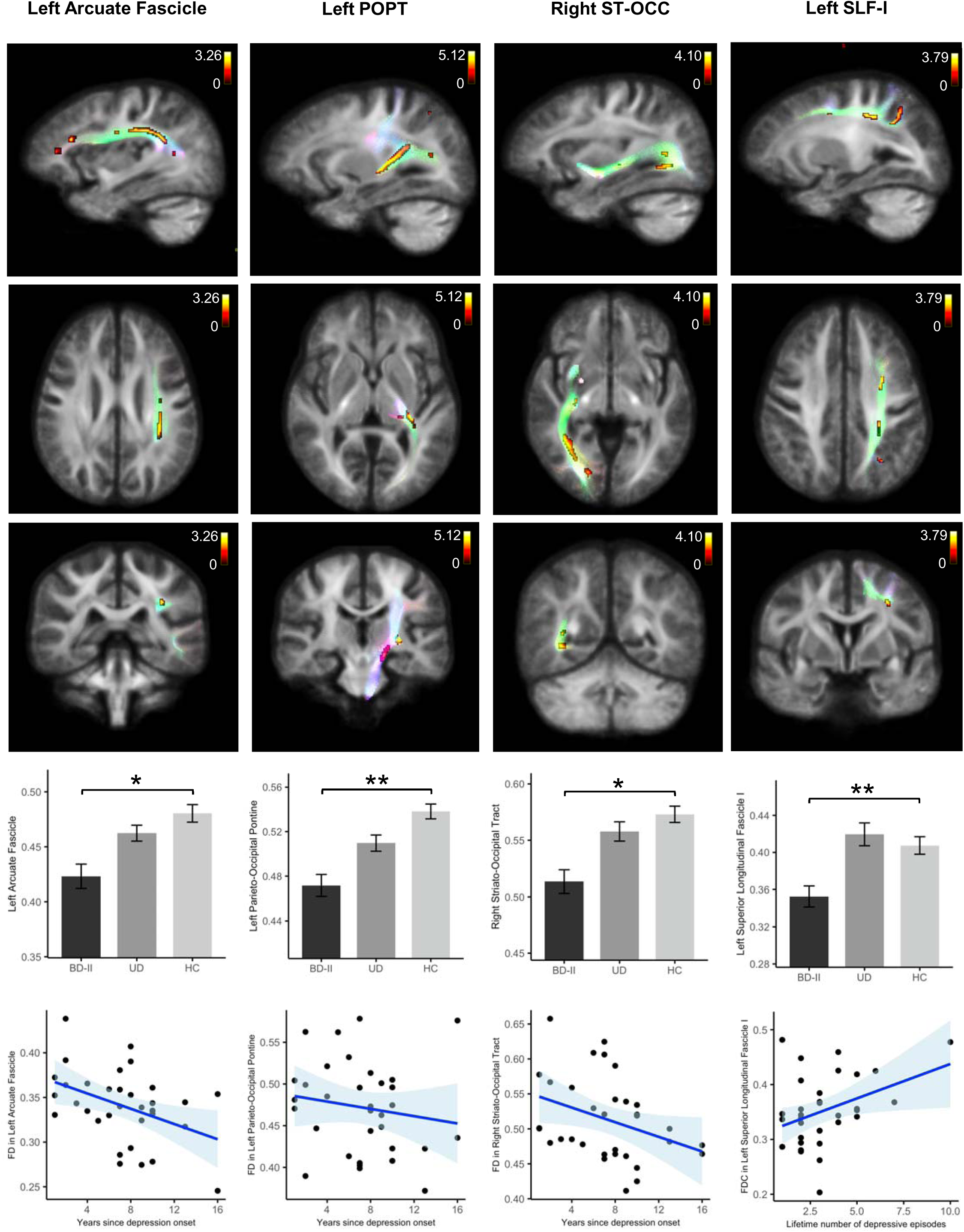

