## Supplementary materials for "White Matter Abnormalities in Bipolar II and Unipolar Depression – Evidence from Fixel-Based Analysis"

S1. **Participant recruitment of the original datasets**

*Pittsburgh dataset*

The Pittsburgh (PITT) dataset belongs to the study entitled “Task preparation abnormality in individuals with unipolar and bipolar depression” by Dr. Anna Manelis of the University of Pittsburgh (hereafter referred to as the “PITT study”).

65 participants were recruited for the PITT study, which included 18 BD-II patients, 23 UD patients, and 24 HC. Participants were recruited from the community, universities, and counselling and medical centres by means of advertisements, referrals, and fliers. Inclusion and exclusion criteria are summarized in Table S1a. All eligible participants provided written informed consent in line with the ethical approval by the University of Pittsburgh and Carnegie Mellon University (CMU) Institutional Review Boards. All experimental procedures were performed in accordance with relevant guidelines.

*HK dataset*

The CUHK dataset was obtained from the study entitled “energy metabolism as biomarker and pathogenesis of bipolar II-concurrent serum measures of lipid peroxidation, 1H-magnetic resonance spectroscopy and diffusion tensor imaging on young treatment-naïve bipolar II and unipolar depression” conducted by Dr. Arthur Dun-ping Mak of the Chinese University of Hong Kong (hereafter referred to as the “CUHK study”).

A total of 81 young adults were recruited for the CUHK study, which included 27 participants in each of the BD-II, UD, and HC groups88. Patients were recruited from individuals presenting at specialist psychiatric clinics for scheduling a new appointment during the years 2014 – 2018, and healthy volunteers were recruited by means of online advertisements and a public health centre. Inclusion and exclusion criteria are summarized in Table S1b.

All eligible participants provided written informed consent in line with the ethical approval (CREC Ref. No.: 2014.168) by the New Territories East Cluster – Chinese University of Hong Kong Clinical Research Ethics Committee. All experimental procedures were performed in accordance with the approved guidelines and regulations.

| **Table S1a. Inclusion and exclusion criteria for recruitment of the PITT study** | | |
| --- | --- | --- |
| **BD-II** | **UD** | **HC** |
| ***Inclusion criteria*** | | |
| Aged above 18 | | |
| Right-handed | | |
| Fluent in English | | |
| Provided written consent | | |
| With current HRSD-25 score > 14 | | / |
| Meeting DSM-5 criteria for BP-II | Meeting DSM-5 criteria for MDD | / |
| ***Exclusion criteria*** | | |
| Meeting criteria for any psychotic-spectrum disorder | | Personal or family history of DSM-5 psychiatric disorders |
| Systematic medical illness | | |
| YMRS score > 10 | | |
| Current alcohol/drug abuse | | |
| Head injury | | |
| Neurodevelopmental disorders | | |
| NART < 85 | | |
| MRI incompatibilities, e.g., metal in the body, pregnancy, claustrophobia | | |
| BD-II, bipolar-II depressed; UD, unipolar depressed; HC, healthy control; HRSD, Hamiltion Rating Scale for Depression; DSM-5, Diagnostic and Statistical Manual of Mental Disorders 5; BPII, bipolar II disorder; MDD, major depressive disorder; YMRS, Young Mania Rating Scale; NART, National Adult Reading Test; MRI, magnetic resonance imaging | | |

| **Table S1b. Inclusion and exclusion criteria for the CUHK study** | | |
| --- | --- | --- |
| **BD-II** | **UD** | **HC** |
| ***Inclusion criteria*** | | |
| Aged 18 - 30 | | |
| Right-handed | | |
| Provided written consent | | |
| Consecutive new referrals to a specialist psychiatric clinic | | / |
| With current MADRS score ≥ 20 | | / |
| Meeting RDC criteria for  BP-II | Meeting DSM-IV criteria for MDD | / |
| ***Exclusion criteria*** | | |
| Any chronic medical condition lasting for more than 3 months | | |
| History of psychotic symptoms | | |
| History of DSM-IV manic episode | | |
| Current or past psychoactive substance use | | |
| Current tobacco use and frequent alcohol use | | |
| Organic brain syndrome or history of brain injury | | |
| Mental retardation | | |
| Current or lifetime history of antidepressant, antipsychotic or mood stabilizer use for more than 2 months | | |
| Any MRI unsafe devices or materials in the body, e.g., pacemakers, cochlear implants and deep-brain stimulators | | |
| / | History of DSM-IV hypomanic episode | Personal or family history of mental disorders |
| BD-II, bipolar-II depressed; UD, unipolar depressed; HC, healthy control; MADRS, Montgomery–Åsberg Depression Rating Scale; RDC, Research Diagnostic Criteria; BP-II, bipolar II disorder; DSM-IV, Diagnostic and Statistical Manual of Mental Disorders IV; MDD, major depressive disorder; MRI, magnetic resonance imaging | | |

S2. **Site-specific demographic and clinical statistics**

| **Table S2a. Demographic and clinical statistics of the Pittsburgh dataset** | | | | | |
| --- | --- | --- | --- | --- | --- |
| **Variable** | **Mean (SD) / Count (%)** | | | **Group difference** | |
|  | **BD-II (n = 18)** | **UD (n = 23)** | **HC (n = 24)** | ***F* / *t* / *X^2^* (df)** | ***p*** |
| Age | 24.7 (4.3) | 25.3 (7.2) | 26.0 (7.1) | 0.2 (2,60) | 0.80 |
| Sex | - | - | - | 0.8 (2.0) | 0.67 |
| *Male* | 5 (27.8%) | 5 (21.7%) | 8 (33.3%) | - | - |
| *Female* | 13 (72.2%) | 18 (78.3%) | 16 (66.7%) | - | - |
| IQ (NART) | 109.4 (5.8) | 112.3 (6.0) | 108.9 (6.7) | 2.0 (2,60) | 0.14 |
| HRSD-17 | 21.0 (4.9) | 16.6 (3.2) | 0.8 (0.8) | 234.7*** (2,59) | < 0.001 |
| YMRS | 3.4 (2.5) | 1.8 (1.6) | 0.4 (0.8) | 15.8*** (2,59) | < 0.001 |
| Family history of BD | - | - | - | 14.1*** (2.0) | < 0.001 |
| *Absent* | 13 (72.2%) | 23 (100.0%) | 24 (100.0%) | - | - |
| *Present* | 5 (27.8%) | 0 (0.0%) | 0 (0.0%) | - | - |
| Lifetime MDE | 5.7 (6.4) | 6.0 (4.5) | - | 0.2 (29.7) | 0.86 |
| Lifetime HME | 2.7 (1.7) | - | - | - | - |
| Age of illness onset | 17.1 (3.4) | 19.0 (7.6) | - | 1.1 (32.0) | 0.27 |
| Illness duration (years) | 8.6 (3.6) | 7.3 (4.5) | - | -1.1 (39.0) | 0.29 |
| BD-II, bipolar-II depressed; UD, unipolar depressed; HC, healthy control; NART, National Adult Reading Test; HRSD, Hamiltion Rating Scale for Depression; YMRS, Young Mania Rating Scale; BD, bipolar disorder; MDE, major depressive episode; HME, hypomanic episode Statistically significant at *p <* (*) 0.05, (**) 0.01, and (***) 0.001 levels | | | | | |

| **Table S2b. Post-hoc pairwise comparison** | | | | | | |
| --- | --- | --- | --- | --- | --- | --- |
| **Variable** | **BD-II – UD** | | **UD – HC** | | **BD-II – HC** | |
|  | ***t* / *X*^2^ (df)** | ***p*** | ***t* / *X*^2^ (df)** | ***p*** | ***t* / *X*^2^ (df)** | ***p*** |
| HRSD-17 | -3.3** (27.6) | 0.002 | 23.2*** (25.0) | < 0.001 | 17.3*** (17.8) | < 0.001 |
| YMRS | -2.3* (27.0) | 0.03 | 4.0*** (31.5) | < 0.001 | 4.9*** (19.3) | < 0.001 |
| Family history of BD | 4.9* (1.0) | 0.03 | - | - | 5.2* (1.0) | 0.02 |
| BD-II, bipolar-II depressed; UD, unipolar depressed; HC, healthy control; HRSD, Hamiltion Rating Scale for Depression; YMRS, Young Mania Rating Scale; BD, bipolar disorder Statistically significant at *p <* (*) 0.05, (**) 0.01, and (***) 0.001 levels | | | | | | |

| **Table S2c. Demographic and clinical statistics of the HK dataset** | | | | | |
| --- | --- | --- | --- | --- | --- |
| **Variable** | **Mean (SD) / Count (%)** | | | **Group difference** | |
|  | **BD-II (n = 27)** | **UD (n = 27)** | **HC (n = 27)** | ***F* / *t* / *X^2^* (df)** | ***p*** |
| Age | 23.6 (4.2) | 24.4 (4.0) | 23.2 (3.3) | 0.7 (2,76) | 0.48 |
| Sex | - | - | - | 1.5 (2.0) | 0.47 |
| *Male* | 6 (22.2%) | 9 (33.3%) | 10 (37.0%) | - | - |
| *Female* | 21 (77.8%) | 18 (66.7%) | 17 (63.0%) | - | - |
| IQ (WAIS-III) | 30.8 (6.3) | 29.1 (7.8) | 35.2 (7.5) | 5.1** (2,76) | 0.01 |
| MADRS | 26.1 (9.2) | 25.6 (5.2) | 0.3 (1.3) | 150.0*** (2,75) | < 0.001 |
| YMRS | 4.2 (4.2) | 1.3 (2.0) | < 0.01 (0.0) | 16.7*** (2,75) | < 0.001 |
| Family history of BD | - | - | - | 18.9*** (2.0) | < 0.001 |
| *Absent* | 16 (59.3%) | 25 (92.6%) | 27 (100.0%) | - | - |
| *Present* | 11 (40.7%) | 2 (7.4%) | 0 (0.0%) | - | - |
| Lifetime MDE | 2.4 (1.3) | 1.4 (0.8) | - | -3.3** (42.1) | 0.002 |
| Lifetime HME | 48.4 (95.1) | - | - | - | - |
| Age of illness onset | 18.7 (5.0) | 21.6 (4.6) | - | 2.2* (51.6) | 0.03 |
| Illness duration (years) | 6.0 (4.1) | 3.9 (3.4) | - | -2.1* (50.6) | 0.04 |
| BD-II, bipolar-II depressed; UD, unipolar depressed; HC, healthy control; Wechsler Adult Intelligence Scale-III; MADRS, Montgomery–Åsberg Depression Rating Scale; YMRS, Young Mania Rating Scale; BD, bipolar disorder; MDE, major depressive episode; HME, hypomanic episode Statistically significant at *p <* (*) 0.05, (**) 0.01, and (***) 0.001 levels | | | | | |

| **Table S2d. Post-hoc pairwise comparison** | | | | | | |
| --- | --- | --- | --- | --- | --- | --- |
| **Variable** | **BD-II – UD** | | **UD – HC** | | **BD-II – HC** | |
|  | ***t* / *X*^2^ (df)** | ***p*** | ***t* / *X*^2^ (df)** | ***p*** | ***t* / *X*^2^ (df)** | ***p*** |
| IQ (WAIS-III) | -0.9 (49.8) | 0.38 | -2.9* (51.9) | 0.02 | -2.3* (50.6) | 0.04 |
| MADRS | -0.3 (41.1) | 0.77 | 24.3*** (29.4) | < 0.001 | 14.4*** (27.1) | 0.003 |
| YMRS | -3.2** (37.0) | 0.003 | 3.5** (26.0) | 0.003 | 5.2*** (26.0) | < 0.001 |
| Family history of BD | 6.5* (1.0) | 0.01 | 0.5 (1.0) | 0.47 | 11.4*** (1.0) | < 0.001 |
| BD-II, bipolar-II depressed; UD, unipolar depressed; HC, healthy control; Wechsler Adult Intelligence Scale-III; MADRS, Montgomery–Åsberg Depression Rating Scale; YMRS, Young Mania Rating Scale; BD, bipolar disorder Statistically significant at *p <* (*) 0.05, (**) 0.01, and (***) 0.001 levels | | | | | | |

S3. **Results of 2(diagnosis)×2(medication) ANOVA**

Table S3 summarizes the results of the two-way ANOVA examining the effects of diagnostic group (BD-II and UD) and medication status on clinical variables in the full dataset. Note that categorical variables (sex and family history of BD) and variable applicable to BD-II only (lifetime number of hypomanic episode) were excluded from this analysis.

Results indicated significant main effects of group on depressive symptoms (*p*=0.02), manic symptoms (YMRS; *p*<0.001), age of illness onset (*p*=0.0096), and illness duration (*p*=0.03). There were also significant main effects of medication status on IQ z-score (*p*=0.007), lifetime number of major depressive episodes (*p*=0.02), and the age of illness onset (*p*=0.03). These results are broadly consistent with the results of one-way ANOVA and t-tests as reported in the body of the manuscript. There were no significant interaction effects between diagnostic group and medication status on any of the clinical variables included in this analysis (*ps*>0.05).

| **Supplementary Table S3. Results of 2(group)** $\boldsymbol{\times}$ **2(medication status) ANOVA** | | | | | | | | | | | | |
| --- | --- | --- | --- | --- | --- | --- | --- | --- | --- | --- | --- | --- |
| **Variable** | **Mean (SD)** | | | | | **ANOVA** | | | | | | |
|  | **BD-II (n = 33)** | | **UD (n = 50)** | | **Group** | | | **Medication status** | | **Group** $\boldsymbol{\times}$ **Medication** | | |
|  | **Medicated (n = 14)** | **Unmedicated (n = 19)** | **Medicated (n = 11)** | **Unmedicated**  **(n = 39)** | ***F*(df)** | | ***p*** | ***F*(df)** | ***p*** | **F(df)** | **p** | |
| IQ (z-score) | 0.6 (0.4) | 0.0 (1.2) | 0.7 (0.4) | -0.3 (1.5) | 0.3 (1,79) | | 0.62 | 7.6** (1,79) | 0.007 | 0.3 (1,79) | | 0.56 |
| Depressive symptoms | 28.8 (7.0) | 26.4 (9.6) | 20.7 (3.8) | 25.0 (5.0) | 5.6* (1,76) | | 0.02 | 0.03 (1,76) | 0.87 | 3.8 (1,76) | | 0.06 |
| YMRS | 3.1 (2.7) | 4.7 (4.4) | 1.3 (1.6) | 1.6 (1.9) | 17.8*** (1,76) | | < 0.001 | 0.3 (1,76) | 0.62 | 0.6 (1,76) | | 0.45 |
| Lifetime MDE | 4.5 (2.1) | 2.3 (1.1) | 4.5 (2.9) | 3.3 (4.1) | 1.1 (1,76) | | 0.31 | 5.3* (1,76) | 0.02 | 1.1 (1,76) | | 0.31 |
| Age of illness onset | 17.1 (3.7) | 18.3 (4.2) | 19.0 (7.1) | 20.8 (6.0) | 7.1** (1,76) | | 0.0096 | 5.1* (1,76) | 0.03 | 0.005 (1,76) | | 0.95 |
| Illness duration (years) | 7.9 (2.7) | 7.3 (5.0) | 6.5 (1.6) | 5.1 (4.7) | 4.9* (1,76) | | 0.03 | 2.4 (1,76) | 0.13 | < 0.001 (1,76) | | 0.98 |
| BD-II, bipolar-II depressed; UD, unipolar depressed; YMRS, Young Mania Rating Scale; MDE, major depressive episode Statistically significant at *p <* (*) 0.05, (**) 0.01, and (***) 0.001 levels | | | | | | | | | | | | |
