## Supplementary material for "White Matter Abnormalities in Bipolar II and Unipolar Depression – Evidence from Fixel-Based Analysis": Figure captions

**Figure 1.**

From top to bottom, statistical maps (row 1: sagittal; row 2: axial; row 3: coronal) and bar plots (row 4) of group differences in fibre density (FD) in, from left to right, the left arcuate fascicle (AF), left parieto-occipito-pontine tract (POPT), and right striato-occipital tract (ST-OCC), and the combined measure of fibre density and cross-section (FDC) in the left superior longitudinal fasciculus I (SLF-I), and scatterplots (row 5) showing the association with clinical characteristics (column 1-3: years since depression onset; column 4: number of lifetime depressive episodes). The statistical maps display the heat map of t value (converted to voxel space) and the TractSeg-derived streamlines of the corresponding tract (green: anterior-posterior; blue: superior-inferior; red: left-right) overlaid on the white matter (WM) fibre orientation distribution (FOD) template.

BD-II, bipolar disorder; UD, unipolar depression; HC, healthy control; POPT, parieto-occipito-pontine tract; ST-OCC, striato-occipital tract; SLF, superior longitudinal fasciculus

Statistically significant at *p* < 0.05 (*), *p* < 0.01 (**)
